# Complete loss of *SLC30A8* in humans improves glucose metabolism and beta cell function

**DOI:** 10.1101/2024.04.05.24305397

**Authors:** Lindsey B. Lamarche, Christopher Koch, Shareef Khalid, Maleeha Zaman, Richard Zessis, Matthew E. Clement, Daniel P. Denning, Allison B. Goldfine, Ali Abbasi, Jennifer L Harrow, Christina Underwood, Kazuhisa Tsunoyama, Makoto Asaumi, Ikuyo Kou, Juan L Rodriguez-Flores, Alan R. Shuldiner, Asif Rasheed, Muhammad Jahanzaib, Muhammad Rehan Mian, Muhammad Bilal Liaqat, Usman Abdulsalam, Riffat Sultana, Anjum Jalal, Muhammad Hamid Saeed, Shahid Abbas, Fazal Rehman Memon, Muhammad Ishaq, Allan M. Gurtan, John E. Dominy, Danish Saleheen

## Abstract

Genetic association studies have demonstrated that partial loss of *SLC30A8* function protects against type 2 diabetes (T2D) in humans, but the impact of complete loss of *SLC30A8* function remains unknown. From whole-exome and genome sequencing of 100,814 participants in the Pakistan Genome Resource, we identified fifteen *SLC30A8* knockouts, including homozygotes for a variant enriched in South Asians (Gln174Ter) and 615 heterozygotes for loss-of-function (LoF) variants. T2D risk was lower in *SLC30A8* LoF hetero- and homozygotes, and the protective effect strengthens in a gene dose-dependent manner (OR_additive_=0.63 [0.53-0.78, p=7.5E-07], OR_recessive_=0.27 [0.09-0.80, p=0.018]). Recall-by-genotype of *SLC30A8* LoF hetero- and homozygotes and their family members with oral glucose tolerance tests showed a gene dose-dependent reduction in glucose levels coupled with elevated insulin. Corrected Insulin Response, Disposition Index, and Insulin Sensitivity Index in LoF hetero- and homozygotes indicated higher glucose-stimulated insulin secretion with preserved beta cell function. These data suggest that therapeutic knockdown of *SLC30A8*, up to and including complete knockout, may treat T2D safely and effectively.

## MAIN

The *SLC30A8* gene encodes the zinc efflux transporter SLC30A8 (ZNT8) in pancreatic beta cells and is one of the strongest type 2 diabetes (T2D)-associated genes to be identified in genome wide-association studies (GWAS)[1-9]. Both common polymorphisms and rare heterozygous loss of function mutations at the *SLC30A8* locus have been associated with reduced risk of developing T2D, a disease characterized primarily by impaired insulin secretion and sensitivity. ZNT8 autoantibodies are also associated with type 1 diabetes[10-12]. Several *in vitro* structure-function studies have proposed mechanisms by which *SLC30A8* LoF directly affects zinc-mediated insulin storage, processing, and secretion[13-24], opening a path to T2D treatment independent of weight loss. While weight loss therapies such as glucagon-like peptide-1 (GLP-1) agonists offer an effective form of treatment for many T2D patients, identification of new targets will benefit patients for whom weight loss drugs are contraindicated and those diagnosed with lean T2D, where the benefit of weight loss therapies is not clear[25]. For this reason, *SLC30A8* has attracted attention in the pharmaceutical industry as a potential target for therapeutic knockdown.

Knockout mouse models have been employed to investigate the effect of *Slc30a8* deletion on metabolic phenotypes such as fasting insulin, blood glucose levels, glucagon secretion, and glucose-stimulated insulin secretion [26-33]. *Slc30a8*-null mice consistently demonstrate lower zinc accumulation and atypical insulin granule formation but show variable effects on insulin secretion and glucose homeostasis depending on the age, sex, diet, and genetic background of the mouse[34-36]. Mouse phenotypes have also shown poor translatability to humans [37].

A 2014 genetic study in Europeans identified two rare protein truncating *SLC30A8* variants, Arg138Ter and Lys34SerfsTer50, for which heterozygosity was associated with 53% and 80% risk reduction for type 2 diabetes, respectively[8]. Despite expansion of the recruitment effort to include ∼150,000 individuals across multiple ancestry groups, no homozygotes of *SLC30A8* LoF variants were identified, limiting the scope of the study to partial loss of *SLC30A8* function. Questions remain about the effect and relative safety of complete loss *SLC30A8* expression in humans.

South Asia is home to approximately two billion people – a large population that is generally under-represented in genetic studies and also disproportionately burdened by diseases like T2D[38]. T2D rates are high and increasing, even for those at lower BMIs[38], making this population particularly relevant for the therapeutic goals of this study. To build upon previous human genetic studies of *SLC30A8*, we use the Pakistan Genomic Resource (PGR), a cohort of Pakistani participants assembled and managed by the Center for Non-Communicable Diseases (CNCD). The PGR is a large biobank enriched in participants from highly consanguineous families. As a result, naturally occurring “human knockouts”, homozygotes for rare loss-of-function variants, are more prevalent than in other biobanks. The PGR consists of 91,512 whole-exome sequences and 9,302 whole genome sequences, for which patient medical history and numerous clinical measurements, including T2D status, are available for most participants. We identify and confirm through whole-exome sequencing both hetero- and homozygotes of for rare protein-truncating and damaging missense variants in *SLC30A8*. For selected variants, we combine *in vitro* protein expression analysis to confirm loss-of-function and phenotypic characterization of human *SLC30A8* knockouts identified through a family-based recall-by-genotype study to evaluate the association between complete *SLC30A8* loss of function with diabetes and other metabolic phenotypes (Figure 1).

**Figure 1:**
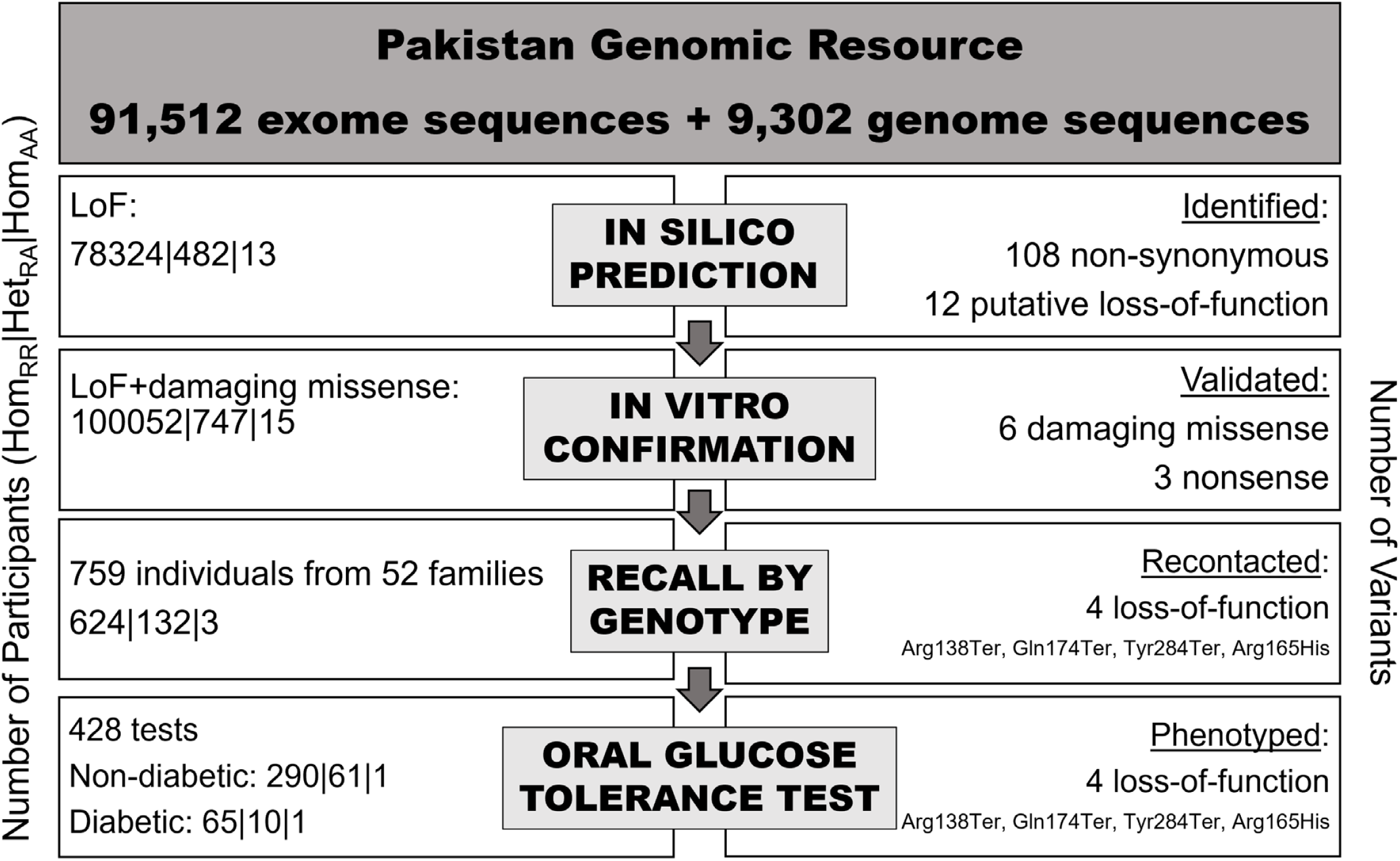
*SLC30A8* variant identification and recall study workflow. All *SLC30A8* coding variants detected in the PGR were first passed through variant effect prediction algorithms to identify putative loss-of-function and damaging missense variants. Selected missense and nonsense variants were profiled via *in vitro* expression testing in HEK293 cells. Probands and their family members were recruited for deep phenotyping studies. These included heterozygotes, homozygotes, and compound heterozygotes for *SLC30A8* loss-of-function variants. Oral glucose tolerance tests were performed when possible.

## RESULTS

### SLC30A8 *is the strongest genetic signal for protection against T2D across the PGR and UKBB*

Genetic associations with T2D in the PGR were identified by ExWAS. To compare the effect of loss of *SLC30A8* function with that of other genes, we conducted an exome-wide gene burden test combining predicted LoF variants or LoF + damaging missense variants with allele frequency below 1%. To boost power, we also conducted a meta-analysis combining the PGR results with the United Kingdom Biobank (UKB) gene burden summary statistics in which similar gene burden definitions were used[39]. In total, our meta-analysis consisted of 33,067 T2D cases and 375,751 controls. A total of six exome-wide (p < 5E-07) significant hits were identified, including *SLC30A8* (p = 5E-08, OR = 0.64 [0.54-0.75]), with consistent effect sizes across the UKB and PGR (p value heterogeneity = 0.83). Of these six genes, *SLC30A8* loss of function was associated with the largest reduction in T2D risk (Fig 2). The only other protective hit for loss of function variants reaching exome-wide significance was *MAP3K15*, which has been reported previously[40].

**Figure 2:**
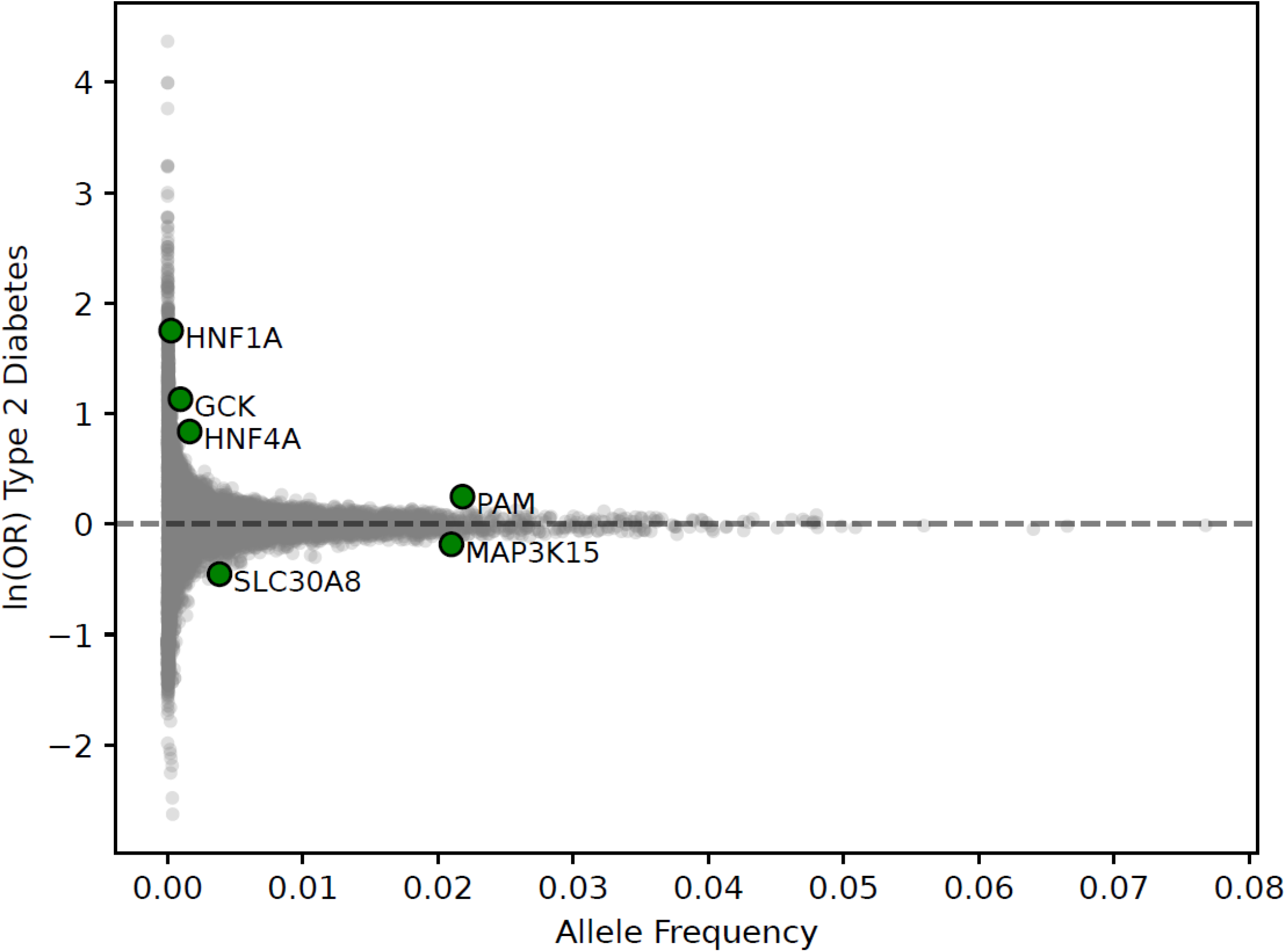
Gene Burden ExWAS for T2D. Significant hits (p < 5E-07) are highlighted. Six genes, *HNF1A, GCK, HNF4A, PAM, MAP3K15* and *SLC30A8* were identified at exome-wide significance. Of the two genes where loss of function was associated with a lower risk of T2D, the greatest reduction in risk was observed for *SLC30A8*.

**Figure 3:**
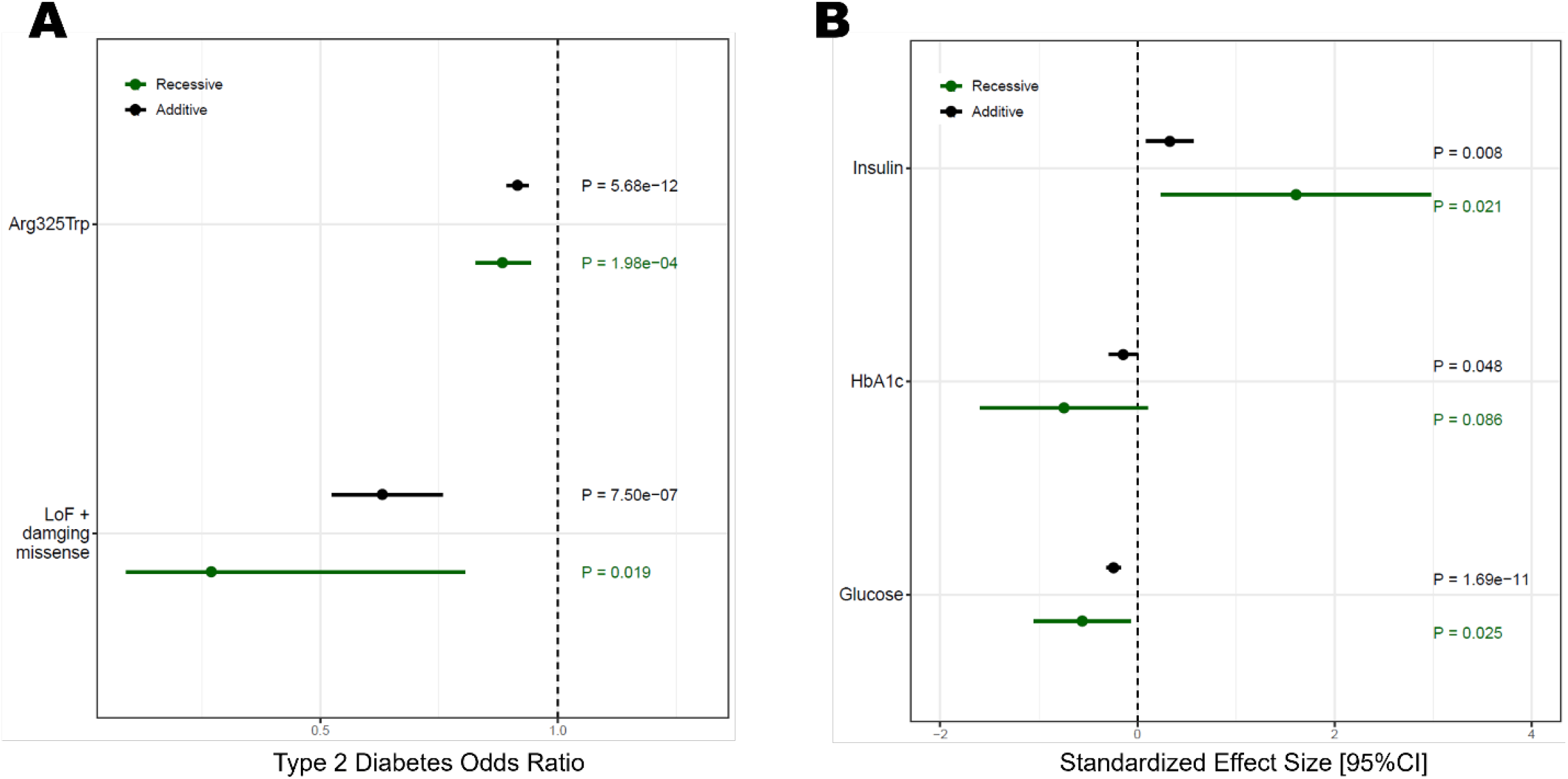
(A) Association with T2D for LoF and damaging missense variants and the common variant Arg325Trp. Additive and recessive models were run adjusting for age, age^2, age*sex, top 10 genetic principal components and whole-genome regression predictions using regenie (B) Association with non-fasting glycemic traits for LoF and damaging missense variants. Traits were rank inverse normalized before analysis. Additive and recessive models were run adjusting for age, age^2, age*sex, top 10 genetic PCs and WGR predictions using regenie

### SLC30A8 *knockouts in the PGR have significantly lower risk for T2D*

Across 100,814 sequenced individuals, we identified sixteen putative LoF variants with a cumulative allele frequency of 0.32% (Supplementary Table 1). These included eleven homozygous individuals and four individuals who were heterozygous for two different nonsense variants. These variants were predicted to be in trans[41], hence these individuals were also included as *SLC30A8* knockouts. Gene burden analysis of LoF hetero- and homozygotes showed a 35% per allele lower T2D risk (OR_additive_=0.66 [0.54 – 0.80], p = 1.6E-05; OR_recessive_ = 0.32[0.09 – 0.98], p = 0.047)

In addition to high-confidence LoF variants, we selected for functional profiling in cell culture six missense variants that were bioinformatically predicted damaging (see methods) or located in highly conserved positions in *SLC30A8* orthologs (Supplementary Fig 1). Both liposome- and cell-based activity assays were attempted to determine variant effects, but no zinc transport activity was observed (data not shown); loss of expression was therefore used to validate damaging missense variants. Five out of six variants showed substantially lower expression of monomeric SLC30A8 and were analyzed for T2D association individually (Supplementary Table 2). Though individual associations were not statistically significant for most variants due to low allele frequency, the Arg165His variant showed a statistically significant reduction in T2D odds ratio (OR_additive_ = 0.50 [0.30-0.84], p = 0.009). We conducted a second gene burden analysis that included the five damaging missense variants in addition to the LoF variants (Supplementary Table 2). In this second analysis, one homozygote of Arg165His and one compound heterozygote of Arg165His and Arg138Ter were included as *SLC30A8* knockouts. Fourteen out of fifteen knockouts did not have T2D, resulting in a 71% decrease in T2D risk (OR_recessive_ = 0.27 [0.09 – 0.80], p = 0.018; OR_additive_= 0.63 [0.53-0.78], p = 7.5E-07). Importantly, the protective effect against T2D was stronger for *SLC30A8* knockouts as compared to heterozygotes.

We also included the common variant Arg325Trp, which has been reported in previous studies to be associated with lower glucose and T2D risk[4, 8-10, 12, 17, 20]. Arg325Trp was also associated with a significant decrease in T2D risk (Supplementary Table 2), however, the effect size was much smaller compared to LoF variants (OR_additive_ = 0.92 [0.89 – 0.94], p = 5.7E-12). When Arg325Trp genotype is treated as a covariate, the observed association between SLC30A8 loss of function and T2D risk reduction remained significant (Supplementary Table 5).

### SLC30A8 *loss of function is associated with lower fasting glucose and higher insulin levels in a gene-dose response pattern*

To explore the effect of *SLC30A8* deficiency, we conducted a burden test of LoF and damaging missense variants for association across a wide range of cardiometabolic and other closely related traits (Supplementary Table 3). We observed significantly lower random blood glucose (β = −0.2 [-0.27, −0.13], p = 1.7E-11), and HbA1c levels (β= −0.08 [-0.22, −0.006], p = 0.048) and higher insulin levels (β = 0.46 [0.21, 0.71], p = 0.008) for *SLC30A8* LoF and missense variants with an additive model. Non-fasted insulin levels were 5-fold higher in knockouts. We also observed a nominal increase in C-peptide levels in knockouts only (p_recessive_ = 0.04). Self-reported family history of diabetes was also significantly lower for heterozygotes and knockouts in an additive model (OR = 0.82 [0.68-0.98], p = 0.03). We observed nominal associations with increased risk of self-reported angina (p = 0.07), higher uric acid (p = 0.003), and higher cystatin C (p = 0.03). No significant associations were observed with other disease endpoints, such as myocardial infarction or other biomarkers including liver enzymes AST, ALT and GGT.

### Recall-by-genotype and oral glucose tolerance test

*SLC30A8* LoF and Arg325Trp probands and their family members were recruited in the largest *SLC30A8* recall-by-genotype study to date. In total, 95 probands were recontacted and 52 agreed to participate (54.7% success rate). For each recontacted participant and their consenting family members, *SLC30A8* genotype was confirmed through Sanger sequencing and measurements of height, weight, heart rate, blood pressure, and lipids were recorded. In total we recruited 132 heterozygotes and 3 homozygotes for LoF variants, as well as 317 heterozygotes and 89 homozygotes for the common variant Arg325Trp for this study.

Oral glucose tolerance tests were conducted for 428 individuals. Serum samples from OGTT participants were analyzed for T2D biomarkers glucose and insulin as well as pro-insulin, C-peptide, and the incretins gastric inhibitory polypeptide (GIP) and GLP-1 (Figure 4A). Results are displayed for 352 non-diabetic participants and fitted with an additive model. We observed an apparent gene dose-response pattern in glucose levels, which were significantly lower for LoF hetero- and homozygotes in the fasted state (β= −0.38, p=5.5E-03) and the first 30 minutes of the test (β= −0.51, p=1.6E-03). We also observed higher insulin levels of LoF hetero- and homozygotes in the first 30 minutes of the test (β= 0.41, p=8.6E-03) and a trend toward elevated insulin levels throughout (β= 0.35, p=4.4E-03). We further analyzed OGTT results for the Arg325Trp variant (Supplementary Figure 2). Compared with LoF hetero- and homozygotes who do not have a copy of the Arg325Trp allele, Arg325Trp hetero- and homozygotes show a similar pattern of lower glucose and higher insulin levels, but with a smaller effect size that did not reach statistical significance.

**Figure 4:**
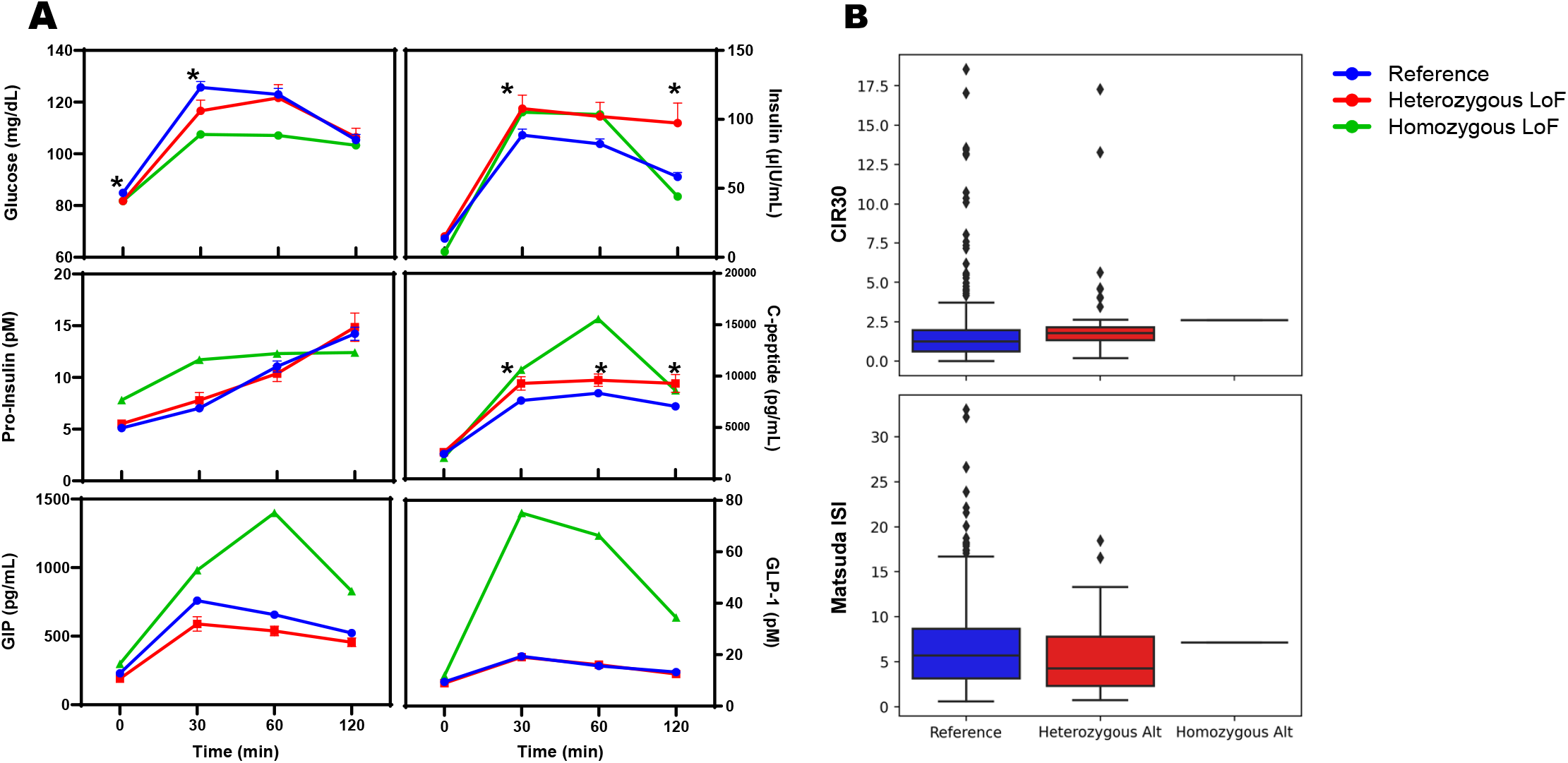
(A) OGTT samples from non-diabetic recall participants were analyzed for plasma glucose, insulin, pro-insulin, C-peptide, GIP and GLP-1. Reference individuals (n=296) do not carry LoF alleles; Heterozygous LoF (n=60) carry one LoF allele; Homozygous LoF (n=1) is a compound heterozygote for two different LoF alleles, Arg138Ter and Gln174Ter; (*) denotes statistical significance (p<0.05). (B) Corrected insulin response (CIR30) and Matsuda index were calculated for each group from relevant insulin and glucose measurements.

From collected OGTT data, Corrected Insulin Response (CIR30), Matsuda Insulin Sensitivity Index, and oral Disposition Index (DI)[42, 43] were calculated as measures of insulin sensitivity and β-cell function in non-diabetic LoF hetero- and homozygotes (Figure 4B). A significantly higher CIR30 (β=0.62, p=1.2E-04) was noted for LoF genotypes, while changes in the Matsuda index did not reach statistical significance (p=0.13). The DI, calculated as a product of insulin response and insulin sensitivity, was also significantly higher for LoF genotypes (β=0.35, p=0.03).

## DISCUSSION

Heterozygous loss of function in *SLC30A8* has been to shown to confer protection from T2D. Leveraging high levels of rare variant homozygosity in PGR, we identified the first complete human knockouts of *SLC30A8*, enabling characterization of loss of function of *SLC30A8* in a gene-dose manner. Using a recall-by-genotype study design, we conducted phenotyping studies in PGR for several metabolic parameters which provided valuable insights into the potential mechanisms of T2D protection conferred by SLC30A8 loss of function.

The PGR is enriched in homozygotes for rare variants because of high levels of parental consanguinity. Initial bioinformatic analyses of PGR participants identified twelve high confidence loss-of-function variants and eight damaging missense variants, of which fourteen are specific to or enriched in South Asia. *In vitro* expression testing confirmed loss of expression for five missense variants, notably Arg165His. In total, twelve homozygous and compound heterozygotes for *SLC30A8* loss-of-function variants were identified, representing the first known human *SLC30A8* ‘knockouts.’ These knockouts included both males and females with an age range of 43-75 years, many of whom have had children and grandchildren. These observations indicate that full *SLC30A8* loss of function from birth is not only compatible with life, but generally well tolerated in humans.

We further conducted deep phenotyping studies on *SLC30A8* LoF hetero- and homozyotes in the largest recall-by-genotype study for *SLC30A8* to date, and the first to include human knockouts of *SLC30A8*. With a study completion rate of 54.7%, this study represents a significant advance over recently reported recall frameworks for T2D[44]. Oral glucose tolerance tests were used as a minimally invasive means to measure glucose dynamics, beta cell function, and insulin resistance. Two identified knockouts were available for OGTT; the first is a diabetic homozygote for the nonsense variant Arg138Ter, the second is a nondiabetic compound heterozygote for both the Arg138Ter and Gln174Ter nonsense variants. Since comparisons of OGTT results are most informative for nondiabetics, only the non-diabetic test is included in our analysis to represent homozygous loss-of-function. The most striking result is significantly lower glucose levels in LoF heterozygotes, coupled with higher post-load insulin levels, in an apparent gene-dose response, indicating better glucose clearance in LoF heterozygotes compared to control group.

From these data, we calculated a CIR30 and Matsuda index to quantify insulin response and sensitivity as a minimally invasive alternative to the traditional hyperglycemic and euglycemic clamp approaches[42, 45]. LoF heterozygotes showed significantly higher CIR30 values than control group, suggesting that improved glucose tolerance is due to enhanced insulin secretion. In contrast, no significant changes were noted in the Matsuda index, suggesting that LoF heterozygotes do not develop insulin resistance. The DI, which approximates beta cell function for the degree of insulin resistance, was also increased in LoF heterozygotes compared to control group, indicating that LoF heterozygotes maintain sufficient beta cell function to compensate for potential insulin resistance. Elevated GIP and GLP-1 were also observed in the non-diabetic *SLC30A8* knockout; however, this pattern was not observed in heterozygotes. More data from additional non-diabetic homozygotes is required to interpret this result. Altogether, these analyses suggest that *SLC30A8* LoF confers protection from T2D through a mechanism of enhanced insulin secretion and improved overall beta cell function, and that the magnitude of this effect may be proportional to the degree of *SLC30A8* loss-of-function.

In summary, we confirm that *SLC30A8* loss of function is protective against type 2 diabetes, with partial loss of function associated with statistically significant improvement in glucose tolerance and insulin secretion. We further show, for the first time, that complete loss of *SLC30A8* function is safe and even well-tolerated in humans, though not completely protective against T2D. Importantly, *SLC30A8* loss of function did not impact BMI (Supplementary Table 3), and therefore represents an alternative mechanism for T2D treatment and prevention independent of currently available weight loss-based therapies. These results provide strong support for *SLC30A8* knockdown as a safe and effective therapeutic approach to the treatment of T2D.

## Data Availability

All academic requests to access relevant data should be sent to. CNCD will ask relevant investigators to sign a data confidentiality agreement which would limit any investigator not to de-identify any of the study participants.

## LIST OF TABLES AND FIGURES

**Supplementary Table 1:**
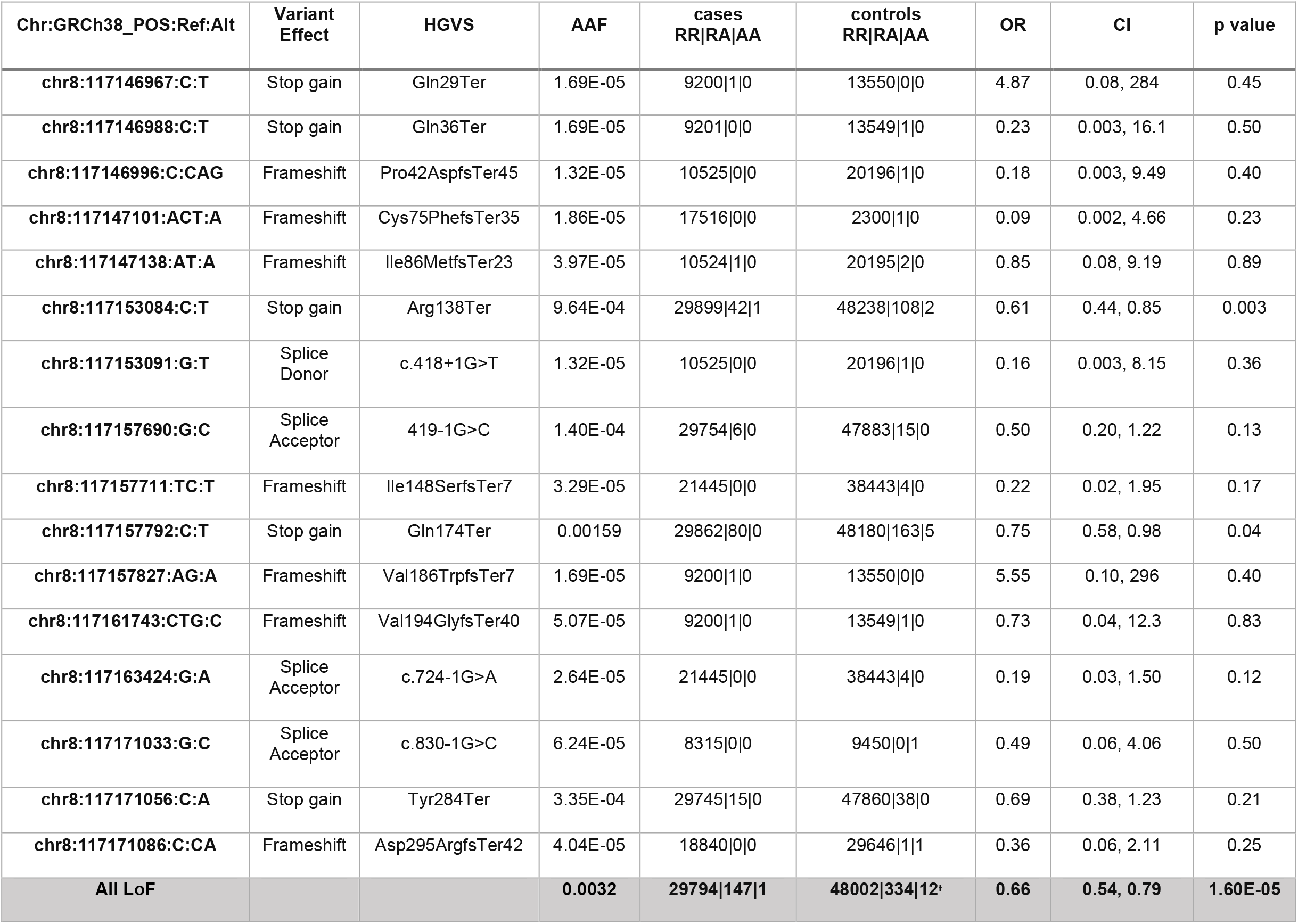
Predicted loss of function variants of *SLC30A8* are shown in a case control analysis for association with T2D individually and in a gene burden test. ^+^One individual was heterozygous for both Arg138Ter and Gln174Ter, and two individuals were heterozygous for both Gln174Ter and Tyr284Ter.

**Supplementary Table 2:** Predicted damaging missense variants of *SLC30A8* are shown in a case-control analysis for association with T2D. Missense variants which led to loss of expression of *SLC30A8 in vitro* were added to the LoF variants from above for the ‘All LoF + damaging missense’ gene burden test. ^+^One individual was heterozygous for Arg165His and Arg138Ter.

| Chr:GRCh38_POS:Ref:Alt | Variant Effect | HGVS | AAF | cases<br>RR\|RA\|AA | controls<br>RR\|RA\|AA | OR | CI | p value |
| --- | --- | --- | --- | --- | --- | --- | --- | --- |
| chr8:117153025:T:C | Missense | Leu118Pro* | 1.50E-04 | 29756\|4\|0 | 47879\|19\|0 | 0.47 | 0.19, 1.14 | 0.09 |
| chr8:117153072:T:A | Missense | Phe134Ile* | 5.50E-05 | 29759\|1\|0 | 47890\|8\|0 | 0.40 | 0.09, 1.78 | 0.23 |
| chr8:117153073:T:G | Missense | Phe134Cys* | 4.98E-05 | 20558\|1\|0 | 34342\|6\|0 | 0.37 | 0.08, 1.81 | 0.23 |
| chr8:117157765:C:T | Missense | Arg165Cys* | 1.39E-04 | 29938\|4\|0 | 483330\|18\|0 | 0.62 | 0.23, 1.67 | 0.35 |
| chr8:117157766:G:A | Missense | Arg165His* | 4.57E-04 | 29929\|13\|0 | 48291\|56\|1 | 0.50 | 0.30, 0.84 | 0.009 |
| chr8:117172544:C:T | Missense | Arg325Trp | 0.283 | 18232\|10104\|1606 | 28146\|17191\|3011 | 0.92 | 0.89, 0.94 | 5.7E-12 |
| chr8:117172652:T:G | Missense | Cys361Gly | 3.38E-05 | 9210\|0\|0 | 13549\|1\|0 | 0.21 | 0.004, 12.5 | 0.45 |
| **All LoF + damaging missense** |  |  | **0.00387** | **29775\|166\|1** | **47914\|420\|14\*** | **0.63** | **0.53, 0.76** | **7.5E-07** |

**Supplementary Table 3:**
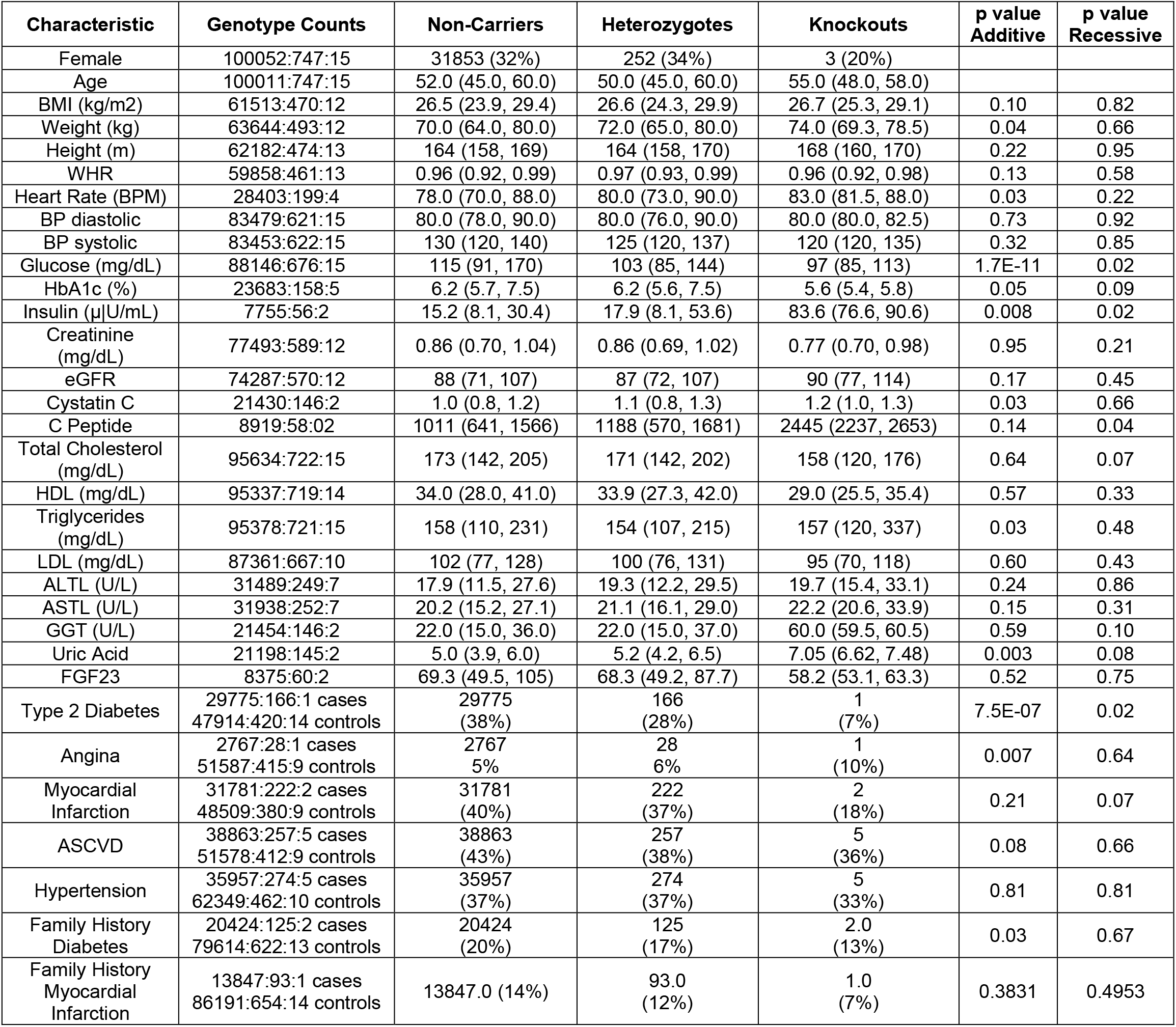
Baseline characteristics and binary traits of *SLC30A8* LoF and damaging missense carriers by genotype. Reported p values were generated by adjusting for age, age^2^, sex, age*sex, top 10 genetic PCs and whole genome regression predictions.

**Supplementary Table 4:** PheWAS of aggregated LoF, missense, and Arg325Trp variants in *SLC30A8* with glycemia related traits. (†) indicates statistical significance, (‡) indicates nominal significance after correction for multiple testing. *SLC30A8* loss of function hetero- and homozygotes demonstrate gene-dose dependent decreases in plasma glucose (**β=** −0.20) and corresponding increases in plasma insulin levels (**β=** 0.46). The effect size is comparable when damaging missense variants are included in the analysis, and remains statistically significant. HbA1c data is limited in this cohort, while the direction of effect is negative as expected for a protective effect, it is not statistically significant in either analysis. A similar but more moderate gene-dose effect is observed for hetero- and homozygotes for the common protective variant Arg325Trp.

| Variants | Phenotype | Genotype Count<br>(Ref:Het:Hom) | Median<br>(Ref) | Median<br>(Het) | Median<br>(Hom) | Beta<br>(per<br>SD) | SE | CI | p value |
| --- | --- | --- | --- | --- | --- | --- | --- | --- | --- |
| **LoF** | Glucose<sup>†</sup><br>(mg/dL) | 88274:550:13 | 115 | 103 | 97 | -0.20 | 0.04 | -0.27-<br>-0.11 | 7.9E-07 |
| | Insulin<sup>‡</sup><br>(μU/mL) | 7768:44:1 | 15.2 | 18.5 | 69.5 | 0.53 | 0.14 | 0.25-0.82 | 0.01 |
|  | HbA1c<br>(%) | 23705:138:3 | 6.2 | 6.2 | 5.8 | -0.07 | 0.08 | -0.23-0.09 | 0.21 |
| **LoF+**<br>**damaging**<br>**missense** | Glucose<sup>†</sup><br>(mg/dL) | 88146:676:15 | 115 | 103 | 97 | -0.20 | 0.04 | -0.27-<br>-0.13 | 2.2E-09 |
| | Insulin<sup>‡</sup><br>(μU/mL) | 7755:56:2 | 15.2 | 17.9 | 83.6 | 0.46 | 0.13 | 0.21-0.71 | 0.02 |
|  | HbA1c<br>(%) | 23683:158:5 | 6.2 | 6.2 | 5.6 | -0.08 | 0.08 | -0.23-0.06 | 0.05 |
| **Arg325Trp** | Glucose<sup>†</sup><br>(mg/dL) | 52537:30994:5306 | 116 | 114 | 112 | -0.04 | 0.01 | -0.05-<br>-0.03 | 1.44E-12 |
| | Insulin<br>(μU/mL) | 4564:2773:476 | 14.9 | 15.7 | 17.2 | 0.01 | 0.02 | -0.02-0.05 | 0.05 |
|  | HbA1c<sup>†</sup><br>(%) | 14103:8276:1467 | 6.2 | 6.2 | 6.1 | -0.05 | 0.01 | -0.07-<br>-0.03 | 9.02E-06 |

**Supplementary Table 5:** Summary statistics comparing association of *SLC30A8* LoF + damaging missense with glycemic traits with and without adjustment for Arg325Trp. Binary trait effect sizes are reported as ln(OR).

|  |  | Not adjusted |  |  |  |  | Adjusted for Arg325Trp |  |  |  |  |
| --- | --- | --- | --- | --- | --- | --- | --- | --- | --- | --- | --- |
| Phenotype | Trait Type | Beta | UCI | LCI | SE | p value | Beta | UCI | LCI | SE | p value |
| Type 2 Diabetes | Binary | -0.46 | -0.29 | -0.64 | 0.09 | 7.5E-07 | -0.48 | -0.30 | -0.66 | 0.09 | 3.0E-07 |
| Family History Diabetes | Binary | -0.20 | -0.016 | -0.38 | 0.09 | 0.033 | -0.20 | -0.023 | -0.39 | 0.09 | 0.027 |
| Glucose | Continuous | -0.24 | -0.17 | -0.31 | 0.04 | 1.7E-11 | -0.25 | -0.18 | -0.32 | 0.04 | 3.6E-12 |
| HbA1c | Continuous | -0.14 | -0.001 | -0.29 | 0.07 | 0.048 | -0.15 | -0.01 | -0.30 | 0.07 | 0.035 |
| Insulin | Continuous | 0.33 | 0.57 | 0.085 | 0.12 | 0.008 | 0.34 | 0.58 | 0.09 | 0.12 | 0.007 |

**Supplementary Figure 1:**
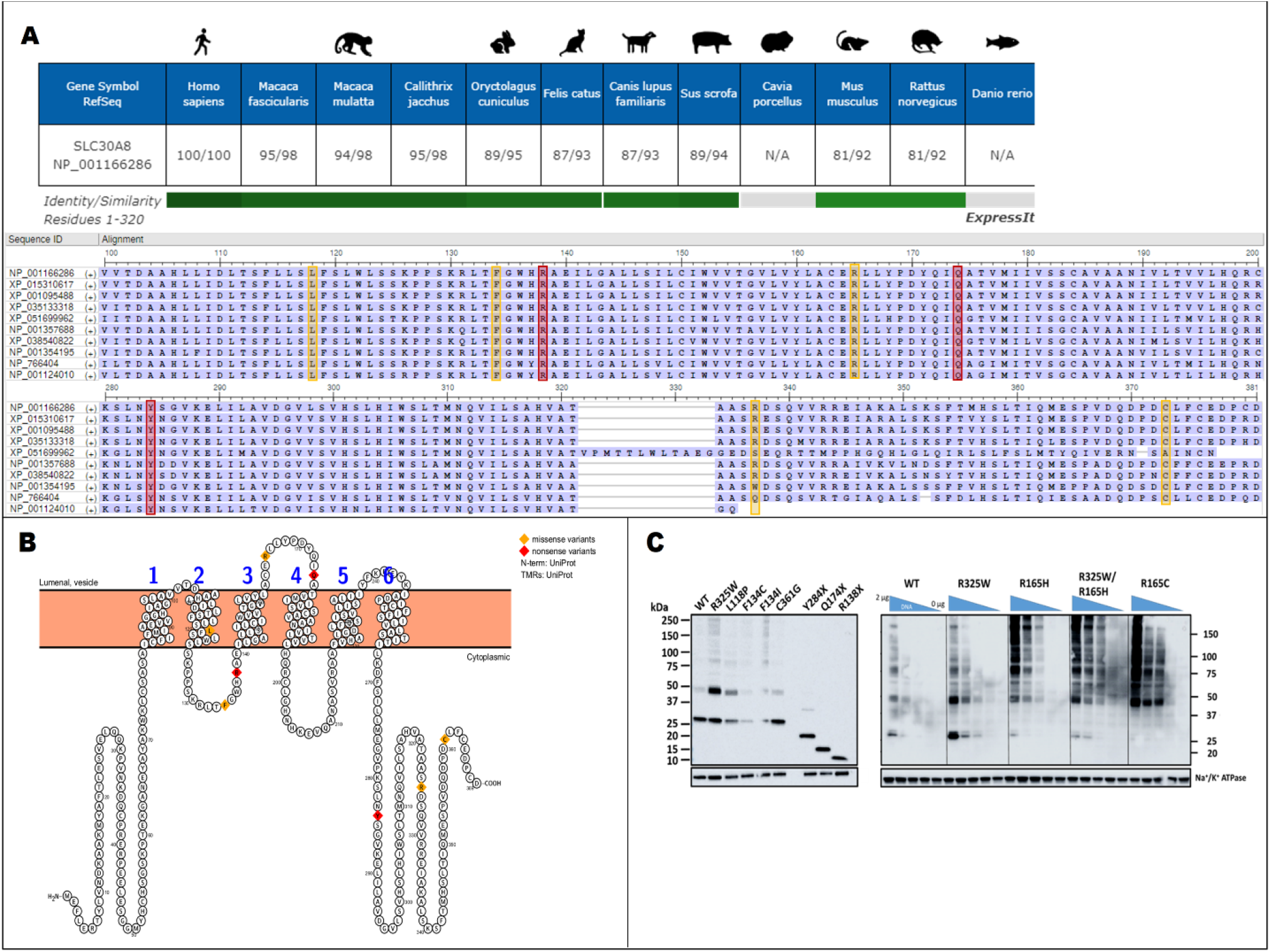
Three nonsense (red boxes) and seven missense (yellow boxes) variants, including Arg325Trp, were selected for *in vitro* expression profiling based on (A) conservation analysis cross *SLC30A8* orthologs or (B) disruption of critical structural features in SLC30A8. (C) cDNA constructs of SLC30A8 variants were expressed in HEK293 cell lysates and detected by western blot against an N-terminal FLAG tag as described previously[24] to allow detection of misfolded and truncated forms of protein which abolish epitopes for SLC30A8-specific antibodies. Monomeric wild-type SLC30A8 (Lane 1) runs as a 30kDa band. Variants that resulted in loss of expression, protein truncation, or significant aggregation relative to wild-type were classified as loss-of-function and included in ExWAS and PheWAS analysis as aggregated “LoF+ damaging missense.”

**Supplementary Figure 2:**
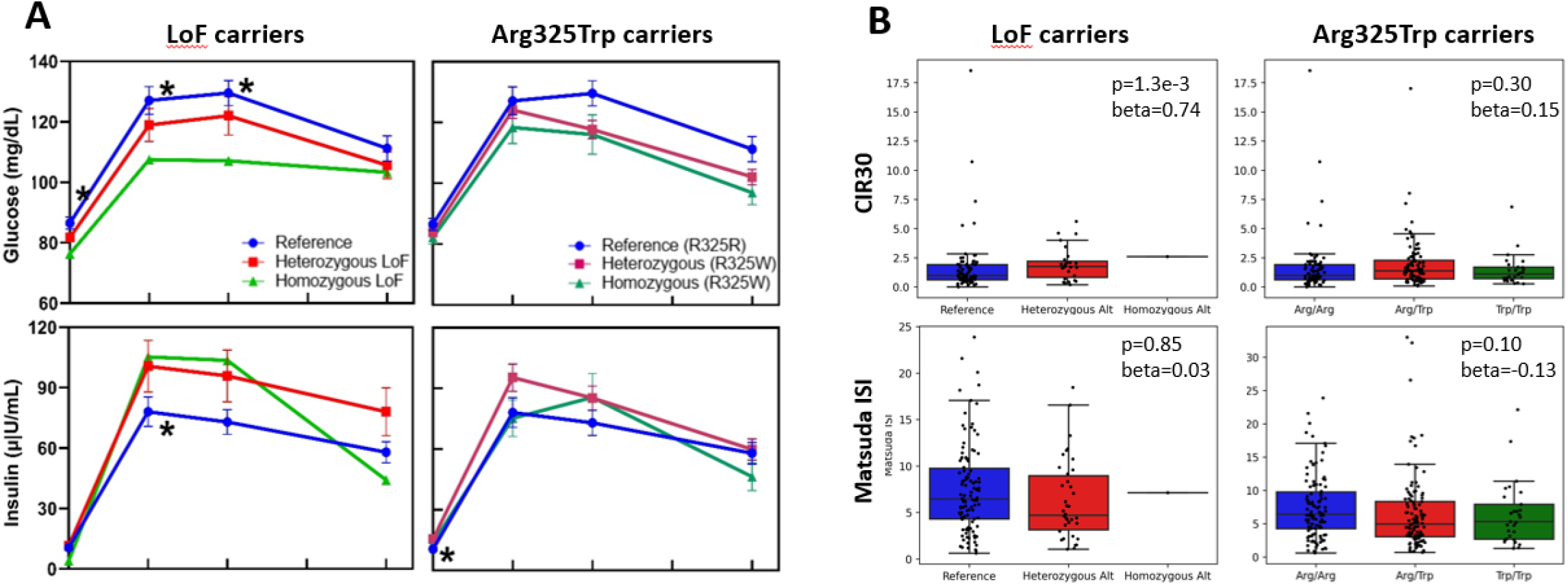
(A) To test for confounding effects of common protective variant Arg325Trp, OGTT results from non-diabetic recall participants were separated by Arg325Trp genotype and re-analyzed. Reference (n=121) do not carry LoF or Arg325Trp alleles; Heterozygous LoF (n=40) carry one LoF allele and no Arg325Trp alleles; Homozygous LoF (n=1) is a compound heterozygote for two LoF alleles and no Arg325Trp alleles; Heterozygous (n=135) and Homozygous (n=33) Arg325Trp carry one or two Arg325Trp alleles, and no LoF alleles. (*) denotes statistical significance (p<0.05). (B) Corrected insulin response (CIR30) and Matsuda Insulin Sensitivity Index calculated for each group from fasting insulin and glucose.

**Supplementary Figure 3:**
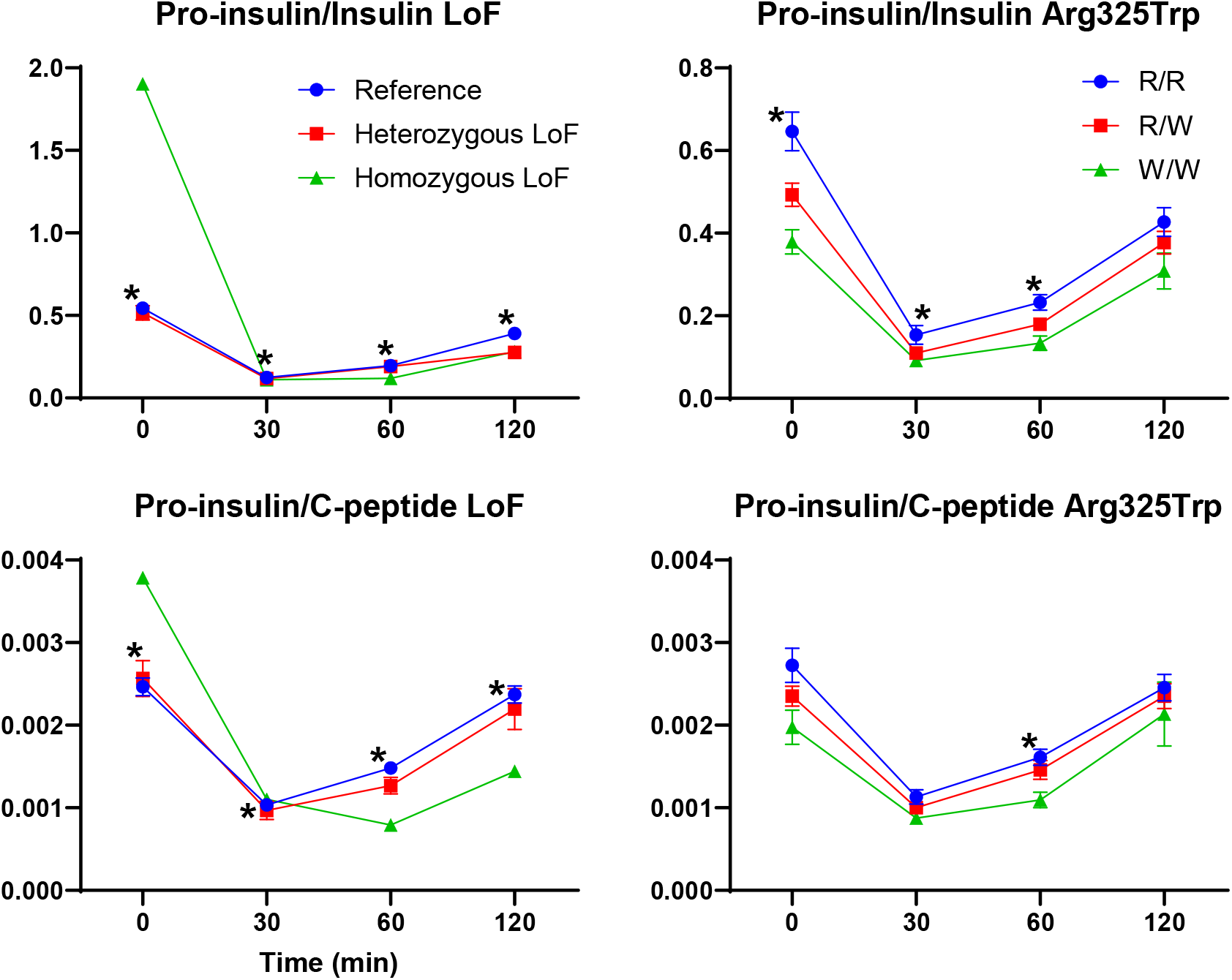
Insulin processing was analyzed with the ratios of pro-insulin to insulin and pro-insulin to C-peptide in OGTT samples. Similar patterns emerged across LoF and Arg325Trp hetero- and homozygotes in which insulin processing efficiency tended to increase slightly in a gene-dose dependent manner. Points marked with (*) represent statistically significant changes (p<0.05).

## METHODS

### Pakistan Genomic Resource Overview

The Pakistan Genomic Resource (PGR) is a collection of nested case control studies comprising of 250,000 participants in Pakistan for whom extensive genetic, biomarker and lifestyle information has been collected enabling evaluation of the genetic determinants of complex diseases and traits. Participants aged 18 or older were enrolled from various hospitals and health units across Pakistan. The Institutional Review Board at the Center of Non-Communicable Disease (NIH registered IRB 00007048) approved the study. All participants provided informed consent. Detailed methodology of PGR is provided in the supplements.

### Phenotype Definitions

Type 2 diabetes was defined as individuals having an HbA1c >= 6.5 or self-reporting to have diabetes or using oral-hypoglycemics. Individuals with a diabetes age of onset less than 30 were excluded from the analysis. T2D controls were defined as individuals who had an Hba1c below 5.7%, or individuals with no self-reported diabetes history or medication. Additionally, individuals with elevated random blood glucose (above 150 mg/dL) were removed as controls[46]. eGFR was calculated using the CKD-EPI calculation. The LDL analysis was subset to individuals who were not on cholesterol lowering drugs, glucose levels were analyzed for participants who were not on oral antidiabetic drugs and creatinine and eGFR were subset to participants without heart failure. Myocardial infarction cases were enrolled at time of event as described[46]. Angina, atrial fibrillation / irregular heartbeats, hypertension were all self-reported. Controls were healthy individuals without any cardiovascular disease history.

### Variant QC and annotation

Exome sequencing was performed at two different locations, at the Broad institute as described earlier and at the Regeneron Genetics Center, as described earlier[47]. All samples are sequenced at 30X coverage. Samples with low allele balance (< 0.2) or low depth (< 10) were set to missing. Male heterozygous call in the non-PAR X chromosome was set to missing and variants that had a missing rate > 5% were removed. High confidence predicted loss-of-function variants were annotated as stop gained, frameshift, splice donor, or splice acceptor variants based on LOFTEE filtering. We used dbNSFP [48] to identify predicted damaging missense variants. Missense variants were categorized as predicted damaging if they attained a REVEL score above 0.8, or if they were flagged as deleterious in 5 out of the 6 following predictors: FATHMM, Sift, PolyPhen, PolyPhen HDIV, LRT and Mutation Taster.

### Exome analysis

All quantitative traits were transformed by the rank based inverse standard normal function, applied within each genotyping batch. Quantitative traits were analyzed using linear regression as implemented in REGENIE. All analyses were adjusted for age, sex, age*sex, age^2^, and top 10 genetics PCs generated using common genotyping array SNPs. For exomes data, if genotyping array data wasn’t available, PCs were derived from common (MAF > 1%) exome SNPs. Exomes and genomes data were analyzed separately across sequencing centers and meta-analyzed using inverse variance weighted meta-analysis as implemented in METAL. Binary traits were analyzed using logistic regression model, with Firth fallback. Additive and Recessive Models were run by specifying the test parameter in regenie.

### Isolation of genomic DNA

DNA was extracted from leukocytes of peripheral whole blood using a reference salting out method. DNA concentrations were determined by UV based quantification by using Nanodrop™ 2000 Spectrophotometer (Thermo Scientific™, USA).

### Sanger sequencing

Genomic DNA from whole blood collected from recalled participants was used to ascertain zygosity for variants of interest via Sanger sequencing. Sanger sequencing was conducted at Macrogen, Inc (South Korea) or in-house at the CNCD. PCR primers were designed covering a region of approximately 200 to 300 bases around the variant. For in-house Sanger sequencing, specific primers were designed to amplify the region of interest using Platinum Master Mix (Thermo Scientific™, USA). This amplified DNA product was cleaned up using ExoSAP-IT Express PCR Product Cleanup (Thermo Scientific™, USA). It was then used for BigDye Terminator v3.1 cycle sequencing following addition of BigDye XTerminator (Thermo Scientific™, USA) for cleanup and run on Applied Biosystems SeqStudio Genetic Analyzer (Thermo Scientific™, USA). Manufacturers’ protocols were followed for all kits.

### Recall-by-genotype

Hetero- and homozygotes for *SLC30A8* LoF variants were contacted by the Center of Non-Communicable Diseases in Karachi Pakistan. After obtaining consent from the proband and from the family members, questionnaires regarding past medical and family history were administered by trained research staff, in the local language. Physical measurements such as height, weight hip and waist circumference were measured in standing position by using height and weight scales. Body mass index was calculated in kg / m^2 units. Waist to hip ratio was measured by taking the standing waist and hip measurements. If standing measures weren’t possible laying waist and hip measurements were taken. Blood pressure and heart rate were recorded by using OMRON healthcare M2 blood pressure monitors. Non-fasting blood samples were collected and analyzed as described previously[47]. A random urine sample was also collected from each participant. The samples were stored temporarily in dry ice in the field and transported to central laboratory based at CNCD and stored at −80°C. Measurements for total-cholesterol, HDL cholesterol, LDL cholesterol, triglycerides, VLDL, AST, ALT and creatinine were conducted in serum samples using enzymatic assays, whereas levels of HbA1c were measured using a turbidimetric assay in whole-blood samples (Roche Diagnostics, USA). Urinary microalbumin were also measured through the urine sample.

### Oral glucose tolerance tests

Consenting participants were requested to be on a high carbohydrate diet three days prior to testing. Blood samples from 8 hour fasting participants were obtained. Participants were then administered glucose (75g, 7.5 oz) orally and 3 mL blood samples were collected at 0, 30, 60 and 120-minute time points. For each sample, plasma and serum were separated, stored on dry ice and transferred to a −80C central location where glucose levels were measured for each time point. Corrected Insulin Response (CIR30) and Matsuda Insulin Sensitivity Index were calculated from fasting glucose and insulin measurements as previously described [42], and the Disposition Index (DI) was calculated as the product of [CIR30xISI]. Analyses were restricted to individuals meeting the following criteria: no reported history of diabetes, HbA1c < 6.5%, and no reported use of glucose lowering medication. Statistical tests were performed using an additive linear mixed model with age, sex, and BMI as fixed effects, and family ID as a random effect. All analyses were performed using the statsmodel Python module (version 0.13.0).

### In vitro expression and western blot

Human codon optimized cDNAs corresponding to wild-type ZNT8B (NCBI ref NP_001166286), missense, and nonsense variants were cloned into pcDNA3.1(+) mammalian expression vector with N-terminal FLAG (DYKDDDDK) tag between the start methionine and second amino acid residues. Transient transfection of adherent HEK293 cells in 6-well plate format was performed with transfection reagent Lipofectamine 2000 (Invitrogen) according to manufacturer’s instructions. Transfection optimization for selected variants was performed by holding constant the DNA to lipofectamine mass ratio at 1:3, and serially diluting *SLC30A8* plasmid DNA into empty vector DNA such that the total plasmid content of each transfection mixture remained fixed at 2 μg while only 2, 1, 0.5, or 0.1 μg of *SLC30A8* DNA was added per well. Cells were harvested 48 hours post-transfection by enzymatic dissociation, washed in PBS, and pelleted by centrifugation at 500xg for 5 minutes. Pellets were resuspended in RIPA buffer supplemented with Halt protease inhibitor cocktail (Thermo Scientific), incubated for 30 min at 4°C with end-over-end rotation, and centrifuged at 16000xg for 15 min to yield solubilized lysate. Lysate samples were analyzed by SDS PAGE and Western Blot against the FLAG epitope to probe ZNT8 expression.

### MSD assays

Circulating levels of insulin, GLP-1 (total), GIP (total), C-peptide and proinsulin were evaluated using multiplexed Meso Scale Discovery (MSD) U-Plex Metabolic Group 1 assay plates according to manufacturer instructions (MSD, catalog no. K151ACL-2). Briefly, 96-well multiplex plates were coated 50 μL per well with biotinylated antibodies coupled to site-specific U-Plex linkers, allowing each analyte to self-assemble on unique spots in each well. Plates were incubated overnight at 4°C with shaking at 500 rpm. Each well was washed three times with 150 μL of wash buffer (provided in kit) before the addition of 50 μL of recombinant protein or human plasma diluted in metabolic assay working solution (provided in kit). Plasma was diluted 8-fold for measurements of GIP, GLP-1, C-peptide, and insulin measurements, or 4-fold for proinsulin measurements. After shaking at 500 rpm for 2 h at room temperature, plates were washed three times with 150 μL wash buffer, treated with 50 μL of detection antibodies conjugated to electrochemiluminescent SULFO-TAG labels per well, and incubated at room temperature for 1hr with shaking. Following three washes with wash buffer,150 μL of MSD-GOLD read buffer B was added to each well, and plates were imaged on a MESO Sector S 600MM Imager (Meso Scale Discovery).The lower limits of quantification (LLOQ) for each analyte were in line with kit specifications (insulin: 0.18 μU/mL; pro-insulin: 1.97 pM; C-peptide: 110 pg/mL; GIP total: 3 pM; and, GLP-1 total: 54.3 pg/mL)

